# Patterns of the COVID19 pandemic spread around the world: exponential vs power laws

**DOI:** 10.1101/2020.03.30.20047274

**Authors:** Natalia L. Komarova, Luis M. Schang, Dominik Wodarz

## Abstract

We have analyzed the COVID19 epidemic data of more than 174 countries (excluding China) in the period between January 22 and March 28, 2020. We found that some countries (such as the US, the UK, and Canada) follow an exponential epidemic growth, while others (like Italy and several other European countries) show a power law like growth. Regardless of the best fitting law, many countries can be shown to follow a common trajectory that is similar to Italy (the epicenter at the time of analysis), but with varying degrees of delay. We found that countries with “younger” epidemics, i.e. countries where the epidemic started more recently, tend to exhibit more exponential like behavior, while countries that were closer behind Italy tend to follow a power law growth. We hypothesize that there is a universal growth pattern of this infection that starts off as exponential and subsequently becomes more power law like. Although it cannot be excluded that this growth pattern is a consequence of social distancing measures, an alternative explanation is that it is an intrinsic epidemic growth law, dictated by a spatially distributed community structure, where the growth in individual highly mixed communities is exponential but the longer term, local geographical spread (in the absence of global mixing) results in a power-law. This is supported by computer simulations of a metapopulation model that gives rise to predictions about the growth dynamics that are consistent with correlations found in the epidemiological data. Therefore, seeing a deviation from straight exponential growth may be a natural progression of the epidemic in each country. On the practical side, this indicates that (i) even in the absence of strict social distancing interventions, exponential growth is not an accurate predictor of longer term infection spread, and (ii) a deviation from exponential spread and a reduction of estimated doubling times do not necessarily indicate successful interventions, which are instead indicated by a transition to a reduced power or by a deviation from power law behavior.

## 1 Introduction

An outbreak of a novel coronavirus, named COVID19, was reported in December 2019 in Wuhan, China, and has been the source of significant morbidity and mortality due to progressive pneumonia [1, 2]. It has since spread around the world and become a pandemic, with large infection burdens reported in Europe, the United States, and in other parts of the world. Disease severity and mortality seem age-dependent, with a higher chance of respiratory complications and death among older people [3], and are further influenced by the availability of health care resources [4]. Non-pharmaceutical interventions, such as social distancing, have been an important tool to slow down the spread of COVD19 [5].

As the non-pharmaceutical interventions are being relaxed in a range of countries, and renewed virus spread is expected to occur, a better understanding of the basic growth dynamics of COVID19 is useful for the interpretation of the emerging epidemiological data. Data from the beginning of the pandemic suggested COVID19 to spread exponentially [6, 7], which is consistent with other epidemics and epidemiological theory [8]. Longer term data, however, suggest that COVID19 spread in China is sub-exponential [9], and it was argued that this is driven by the implementation of strong non-pharmaceutical interventions. A more comprehensive analysis of the growth dynamics of this infection before strict lockdown measures were put in place, however, remains to be carried out. In this study, we compare the per capita virus spread kinetics observed for many countries around the globe during the time frame before strict interventions were put in place, in order to obtain a better understanding of similarities and differences. While some countries can be better described by exponential growth, many other countries are more accurately described by a power law. Interestingly, we find that the growth dynamics become more power-law like if the epidemic is more advanced. This indicates that the long-term dynamics of COVID19 spread might be intrinsically governed by a power law, even in the absence of strict non-pharmaceutical interventions. We interpret these findings with computer simulations of a metapopulation model, which can account for an initial exponential spread phase, followed by a longer-term power law behavior. We relate model predictions to epidemiological correlations found in the data. Because power law growth results in a slow-down of the infection growth rate over time even in the absence of strict interventions, these insights are important for the assessment of the developing pandemic and of the effectiveness of non-pharmaceutical interventions.

## 2 Data sources

The data of confirmed COVID19 cases over time have been obtained from the data repository maintained by Johns Hopkins University Center for Systems Science and Engineering (CSSE) [10]. As of March 28, 174 countries were represented in the database, as well as the cases on “Diamond Princess” (which were not used in the analysis). We only included the total counts for each country, even though information on the different provinces was available for several countries. The number of confirmed cases has been recorded since January 22, 2020, and has been updated daily.

We also used “Our World in Data” [11] to collect information on the daily number of tests, the number of positive tests, and the number of deaths in different countries.

To compare the time course of COVID19 cases across different locations, the per capita incidence was calculated, normalizing the numbers by the total population size of the country. The information on the population size and the area of different countries was taken from Wolfram Mathematica’s database, “CountryData”. Challenges arising as a result of differences in testing policies in different countries and in the same country over time are discussed below.

The COVID19 data analyzed here span the period until March 28, 2020. Soon after that date, many countries experienced a significant slowing down and “saturation” in the infection spread, which indicates the influence of factors that go beyond the scope of the present paper, where we seek to understand the basic laws of initial infection spread.

It is important to note that while the Johns Hopkins data set is comprehensive and is based on many data sources, including government data, it is possible that they contain inaccuracies, e.g. data might not be backfilled if they have an earlier onset date. The supplement compares the Johns Hopkins data to those presented by the Italian health ministry, and we find good agreement. The exact methodology used by the Johns Hopkins data source is provided in [12], which makes clear the advantages and disadvantages of this dataset.

## 3 Results

### 3.1 Per capita case numbers and time lags

Here, we present the comparison of the kinetics according to which cumulative COVID19 cases grow over time in different countries around the world. Figure 1(a) presents the raw data showing total case counts for a select number of countries. Figure 1(b) shows the corresponding per million case counts.

**Figure 1:**
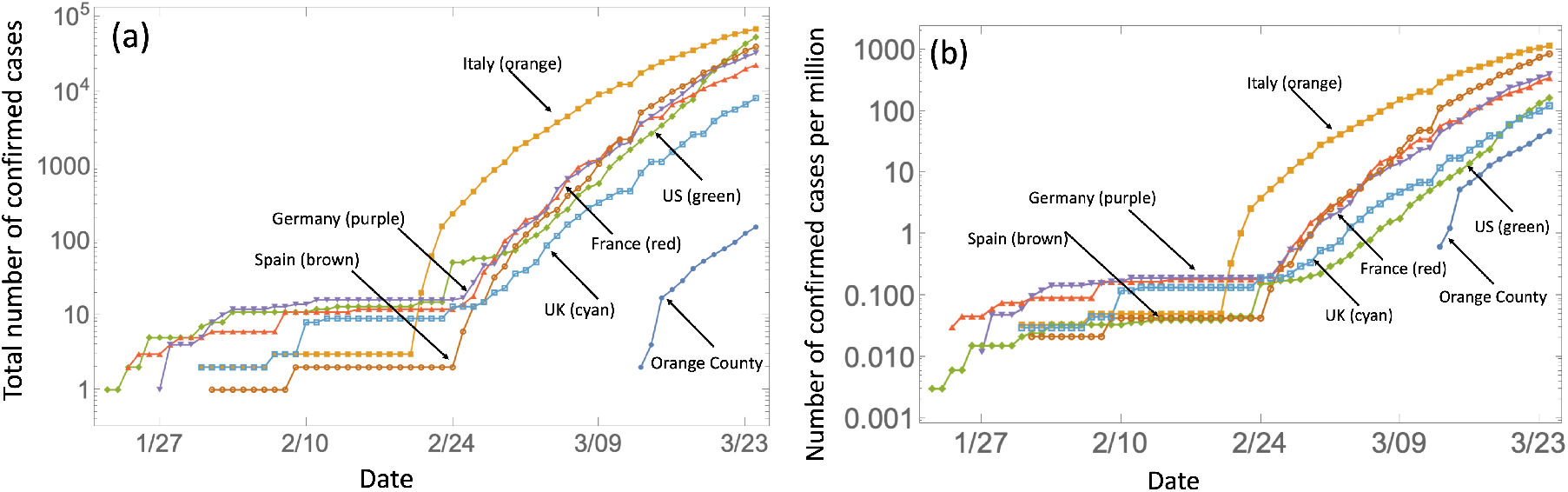
Example of the data. The number of confirmed cases is plotted as a function of time for 6 countries and Orange County: (a) the raw counts, (b) cases per million. The numbers of confirmed COVID19 cases in Orange County, the home of the authors, have been obtained from the daily updates provided by the website of the Orange County Health Care Agency (OCHCA).

A complication for comparing the growth dynamics is that the timing of the onset of community spread varies across locations. The growth curve of confirmed cases was therefore shifted in time to make them comparable, according to the following procedure. The cumulative confirmed COVID19 case counts in Italy were chosen to be the example against which the growth curves in all other countries were compared, due to Italy being an epicenter of the outbreak at the time of this analysis. The (normalized, cases per million) infection growth curves of the other locations were shifted in time such that the difference between all data points of the country under consideration and Italy was minimized. The shift that minimized this Euclidean distance between the curves was assumed to indicate the number of days that the country lags behind Italy. Some examples of such results are presented in figure 2. We note that this assumes that all the countries test for COVID19 at comparable levels, which is an over-simplification. If a country tests less than Italy, it will lag behind Italy to a lesser extent. Conversely, if a country tests more than Italy, it is predicted to be further behind Italy. A more in depth description of the role of testing is given in the “Discussion and Conclusion” section.

**Figure 2:**
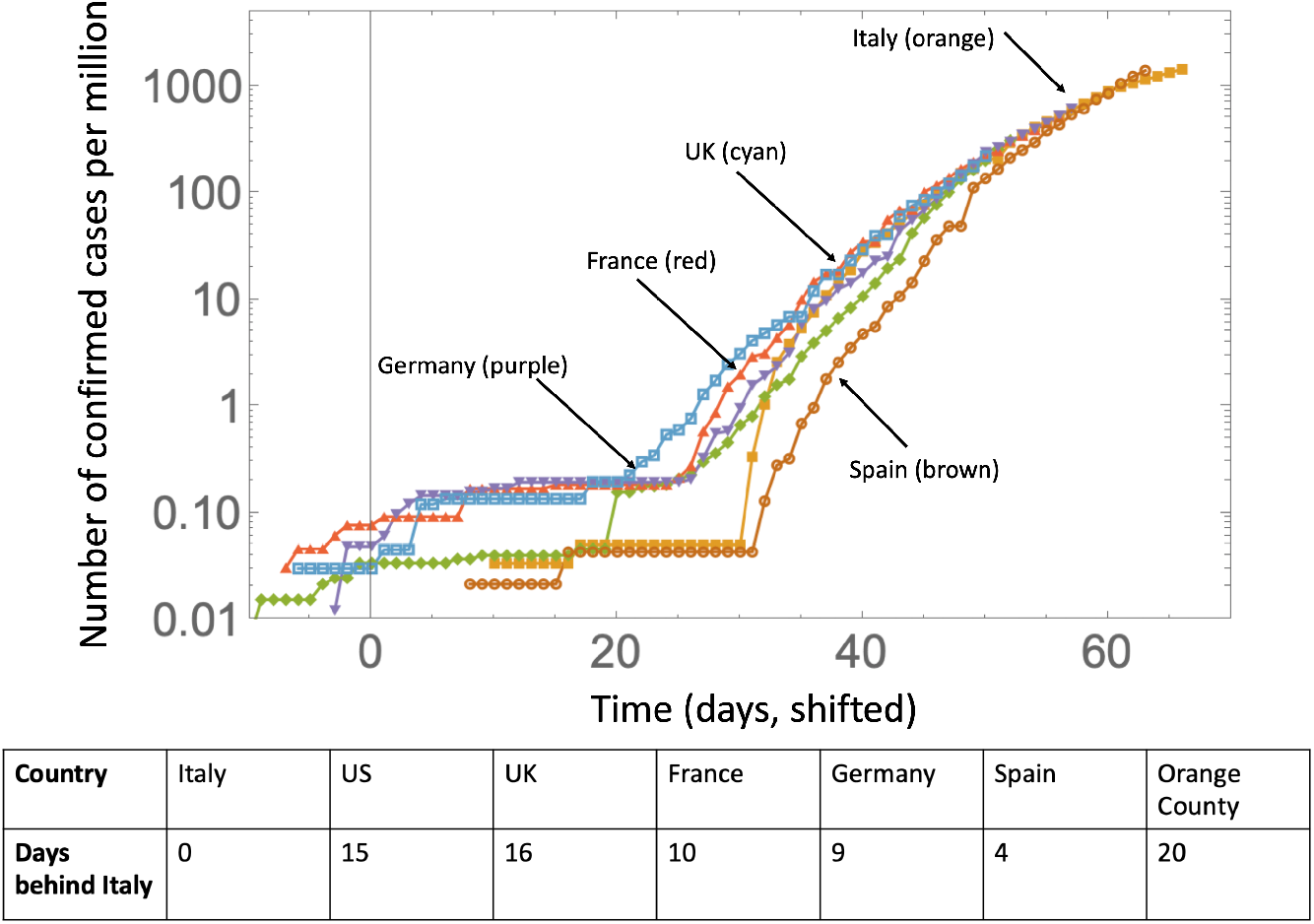
The same data as in figure 1(bottom), presented by shifting individual lines to match the Italy curve. The table shows the lag, that is, by how many days each country is behind Italy.

In this way, we obtained a time course of confirmed COVID19 cases that are temporally synchronized with Italy, which allows for a more straightforward comparison of the kinetics. An interesting observation in figure 2 is that over time, the different countries converge to a similar, sub-exponential growth pattern. While the onset of a certain degree of social distancing could account for sub-exponential growth [9], the observed similarity in the growth patterns across the different countries might argue against this explanation. We therefore hypothesize that the sub-exponential growth patterns are an intrinsic characteristic of COVID19 spread. We explore this hypothesis in detail in the following sections.

### 3.2 Growth laws of the epidemic in different countries

The above analysis indicated that COVID19 spread in a number of countries is sub-exponential. Previous work [13] has suggested that a power law might be a good description of the cumulative COVID19 cases over time in China during the earlier stages of the pandemic. Therefore, we hypothesized that for a subset of the countries, a power law is an appropriate description. To test this hypothesis, we fit both exponential and power law curves to the data for each country and determined the goodness of fit.

Data fitting was performed as follows. Only the data points were considered where the number of COVID19 cases had risen above a threshold, which we set at 1 case per million people (see Supplement for variations of this threshold). We fit both a power function and an exponential function to the data to determine which model fits the data better. For the power law function, a complication arose because fitting requires knowledge of the “zero” point, that is, the moment of time when the growth (according to the power law) began. The fits to the data change if the time scale is changed. Hence, we started by assuming the first data point to be the day when the infection frequency first exceeded 1 case per million, and fitted the power law, *a_1_x^b1^*, for some constants *a_1_* and *b*_1_. Then we shifted the time series incrementally by one day, and for each shift the power law was fitted. For each fitting frame, a different value of the power law exponent, *b_1_*, was obtained. Subsequently, we fit an exponential function to the same data (*a_2_e^b2x^*). The estimated exponent does not depend on the time shift, so fitting the exponential function was straightforward and yielded a unique value b2 for all the fitting frames. For both the exponential and the power law fits, we determined the sum squared error between observed and expected, and compared them. We also used the Aikaike Information Criterion to distinguish between exponential and power law fits and found results to remain robust (see Supplemental Section A).

Figure 3 shows the fitting errors calculated for 75 countries; we included a country if the number of cases reached 20 per million, and excluded China and South Korea, since their epidemics clearly deviate from an exponential or a power law. Since in smaller countries (such as for example Luxembourg) the laws may be harder to determine and the data are subject to a higher degree of noise, for classification purposes we restricted the pool of countries to those with over a million inhabitants. The yellow horizontal lines in figure 3 represent the exponential fitting and the blue lines the power law fitting, as a function of the frame shift. We observe that there are several different configurations that are repeatedly encountered.

**Figure 3:**
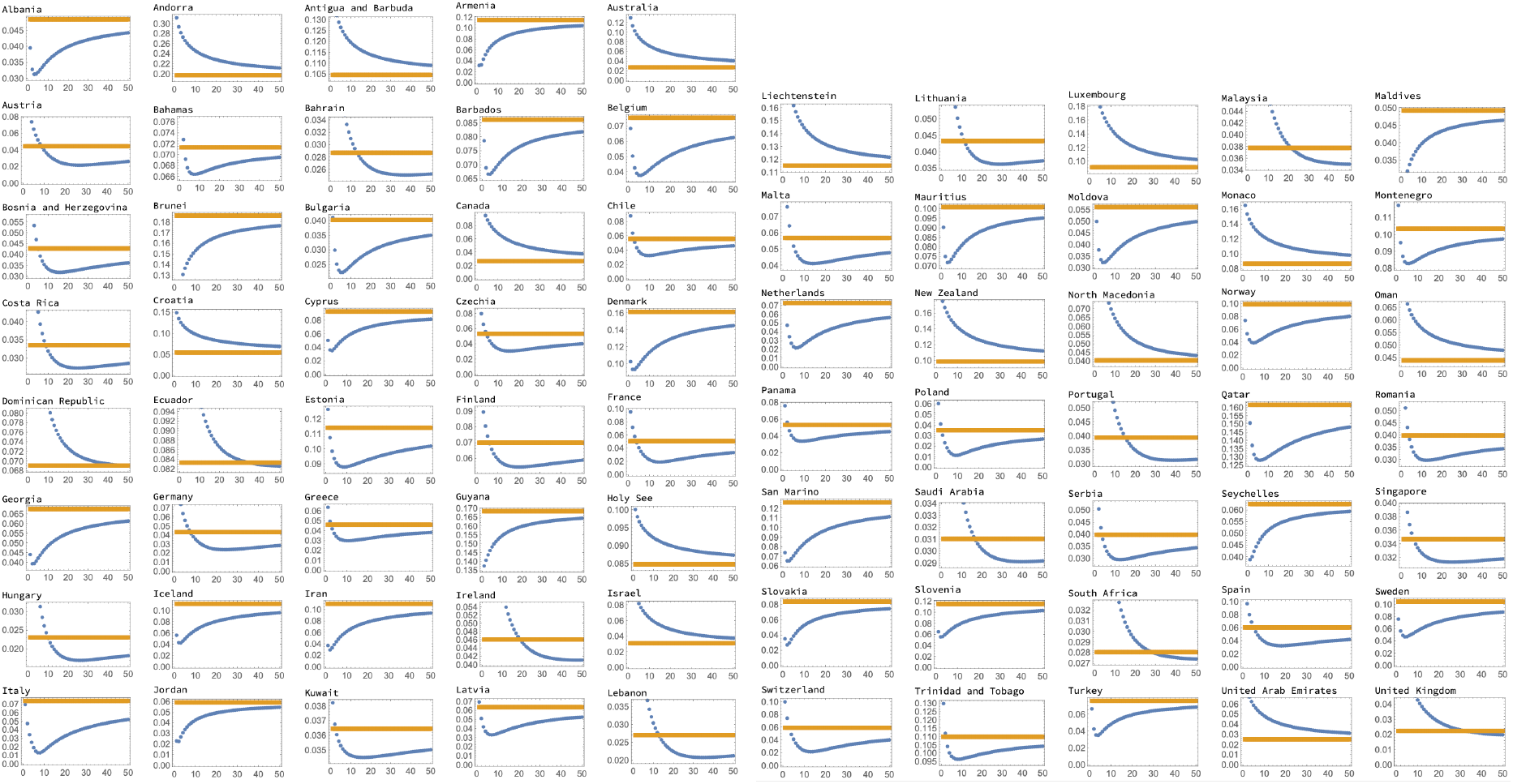
75 countries’ fitting results are presented as errors (blue for power law fits and yellow for exponential fits) as functions of the frame shift. Three distinct configurations can be observed: blue below yellow (a clear power law case), blue above yellow (a clear exponential case), and blue intersecting yellow. For such intermediate cases, we classified the growth as power-like if the power corresponding to the point of intersection corresponded to the power *b*_1_ < 5. Otherwise it was classified as exponent-like.

- For some countries (like the US, see also figure 4(a)), the power fitting error is always above the exponential fitting error. Such countries are clearly showing an exponential epidemic growth.
- There is another group of countries (such as Italy, see also figure 4(b)), where the power law fitting error is always below the exponential error; here we clearly have a power law growth.
- There are some other countries that we can classify as power law-like and exponential-like. Suppose a power law error curve crosses the exponential error line (see Greece, figure 4(c)), at a given frame shift. In this case, we will classify the growth as power law-like if the value of the exponent *b*_1_ that corresponds to this frame shift satisfies *b*_1_ < 5. Otherwise, we will classify the growth law as exponential-like.

**Figure 4:**
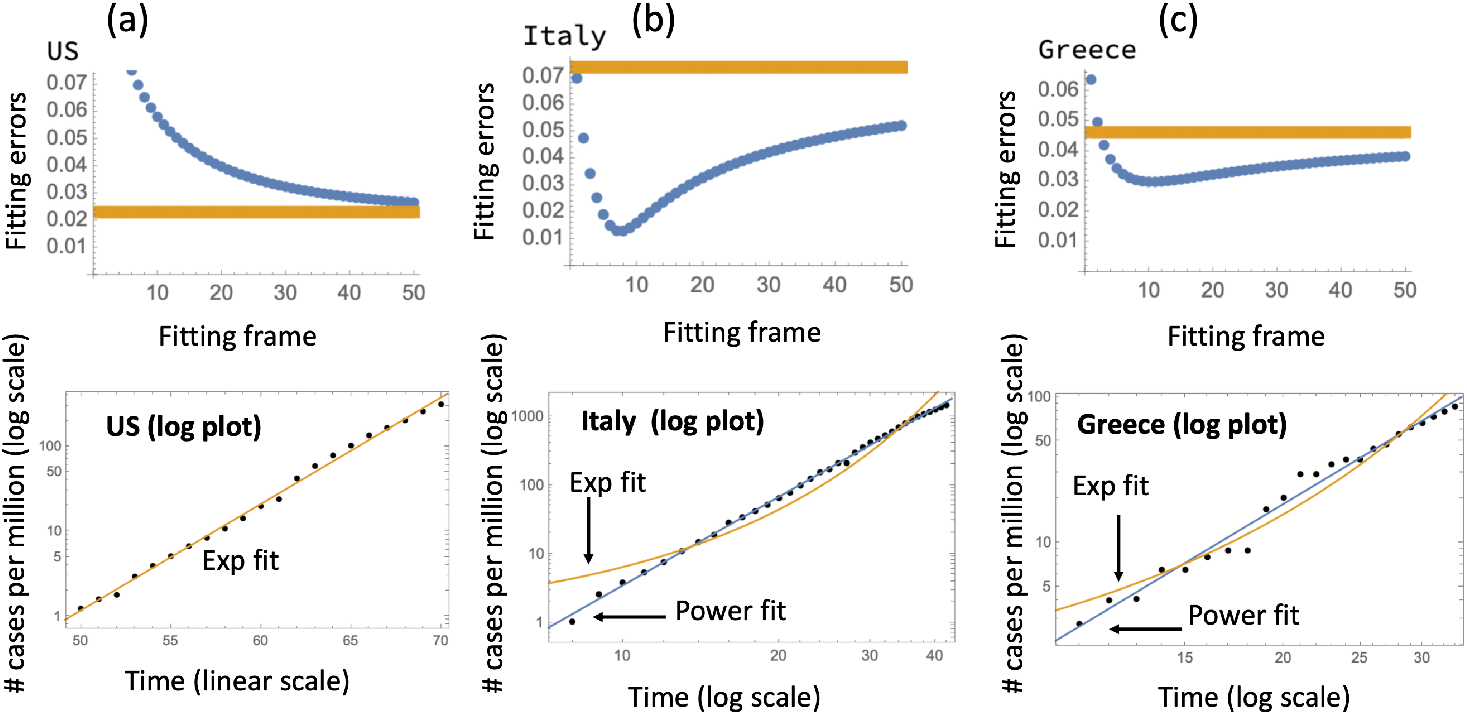
Examples of three error graph configurations. (a) US, exponential; a log plot of the data is presented with the exponential fit. (b) Italy, power-law; a log log plot of the data is presented with the best fitting power law and exponential fits. (c) Greece, exponential-like; as in (b), a log-log plot is presented.

For the examples mentioned here, figure 4 shows the best fits obtained by this method. For (b) and (c) it is clear that the power law provides more satisfactory fits. More details are provided in the Supplement.

A classification of all the countries into those that follow power law (or power law-like) dynamics and those that follow exponential (or exponential-like) growth is given in table 1. About 70% of the countries included in this analysis were classified as displaying power law (or power law-like) dynamics, indicating that this is a wide-spread phenomenon around the world. Geographic distribution of the countries with different growth laws is shown in figure 5.

**Table 1:** Classification of countries according to the epidemic growth law.

| Law | # | List of countries |
| --- | --- | --- |
| Exponential | 9 | Australia, Canada, Croatia, Israel, New Zealand, North Macedonia, Oman, United Arab Emirates, US |
| Exponential-like | 9 | Austria, Dominican Republic, Ecuador, Ireland, Lithuania, Malaysia, Portugal, South Africa, United Kingdom |
| Power law | 20 | Albania, Armenia, Belgium, Cyprus, Denmark, Georgia, Iran, Italy, Jordan, Mauritius, Moldova, Netherlands, Norway, Qatar, Slovakia, Slovenia, Sweden, Turkey, Uruguay |
| Power law-like | 23 | Bahrain, Bosnia and Herzegovina, Bulgaria, Chile, Costa Rica, Czech Republic, Estonia, Finland, France, Germany, Greece, Hungary, Kuwait, Latvia, Lebanon, Panama, Poland, Romania, Saudi Arabia, Serbia, Singapore, Spain, Switzerland, Trinidad and Tobago |

**Figure 5:**
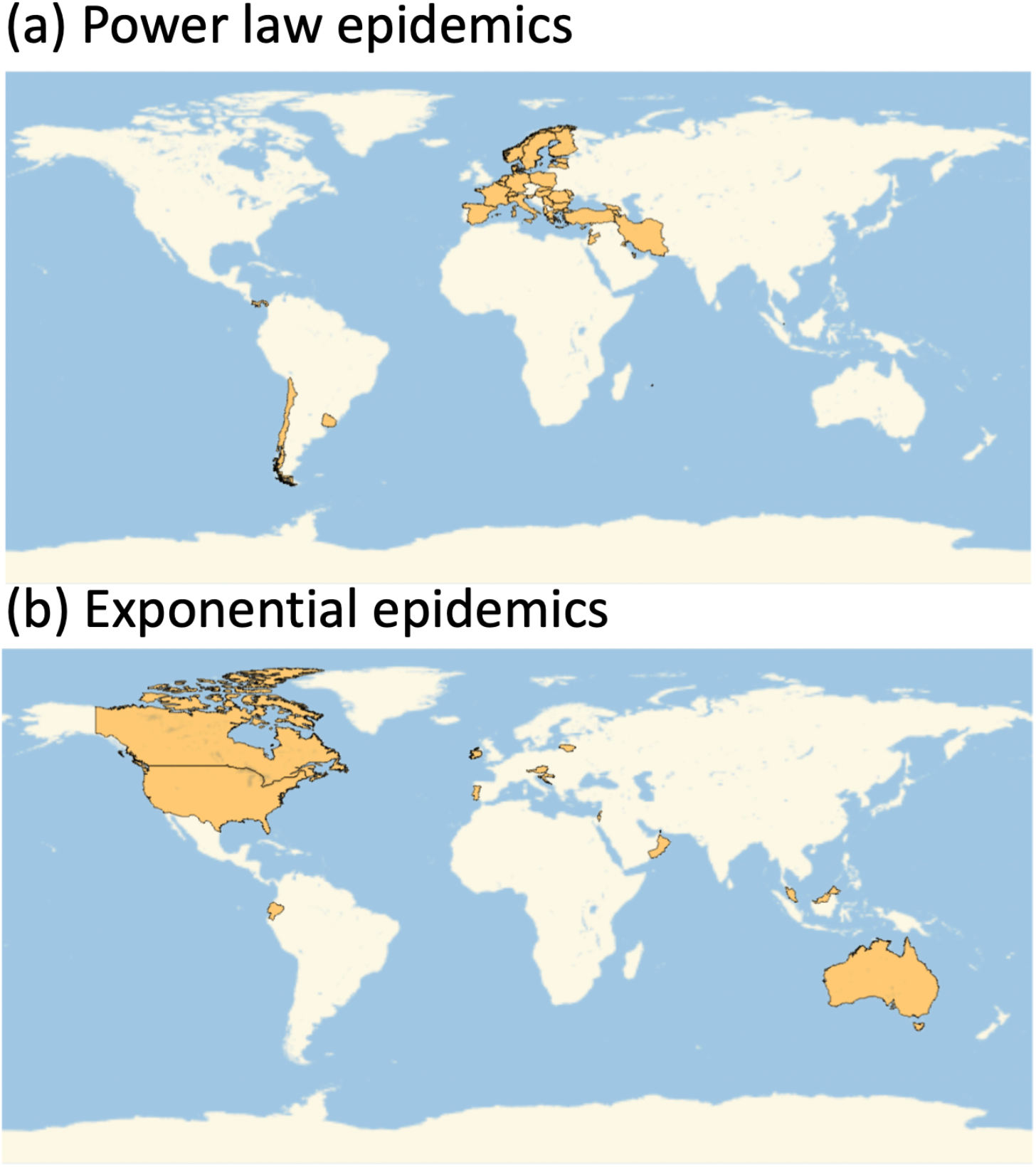
Geographic distributions of different epidemic growth laws. (a) Power law epidemics; (b) Exponential epidemics.

We note that growth laws could potentially be impacted by changes in the level of testing over time in a given country. We therefore have analyzed the dynamics of testing (Supplemental Section C) and saw that the number of tests has typically increased in most countries. This means that the infection growth curves for those countries are accelerated compared to the “true” epidemic growth curves. Thus, if a country is classified as a “power law” or “power law-like” by our analysis, it is unlikely that correction for an increase in testing would move it to an “exponential” or “exponential-like” category. On the contrary, it is possible that some of the epidemic curves that were classified as exponential are in fact slower growing. This makes our list of “power law” and “power law-like” countries a conservative list, which may potentially be larger due to effects of increasing testing. Furthermore, we have performed an analysis of death data (Supplemental Section D). For countries on the list that provided death data during the time-frame of the analysis, we found that a large majority have a power law like death curve, and this majority is larger among the countries in table 1 that we classified as “power law (like)” compared to “exponential (like)”, which also corroborates our findings.

### 3.3 Growth laws in relation to the local stage of the outbreak

We investigate how the growth law of COVID19 spread correlates with the stage of the outbreak in the different countries (i.e. how “old” the outbreak is). Figure 6(a) shows a numerical probability distribution for the day when the infection in each country reached the level of 1 case per million. If this mile stone is reached earlier in a given country, the spread is more advanced. Blue represents the power law set and yellow the exponential set (grey means an overlap of the two colors). The average date of reaching 1 case per million (counting from Jan 22) is about 48 days for the power law and 52 days for the exponential set (*p* = 0.035 by T test). This means that the countries with a power law spread were at a slightly more advanced stage of the epidemic than the exponentially developing countries. This points us towards a hypothesis that perhaps it is typical to observe a transition between an early, exponential stage of growth, and a later, power-like stage of growth. In other words, different countries are at different stages of epidemic development, but they all roughly follow the same trajectory, where an initial exponential growth is gradually replaced by a more power like behavior. Figure 7 demonstrates further evidence in favor of this theory. Panel (a) plots the number of countries classified by the number of days they are delayed with respect to Italy. As explained in section 3.1 (see also figure 2), we shifted the growth trajectories of all countries until, for each country, the best match with the Italian curve was obtained. As we can see in figure 7(a), there are only a few countries that are just behind Italy, and as the number of lag days increases, the number of countries grows. This corresponds to more and more countries becoming affected as time goes by. Figure 7(b) calculates, for each lag time, the percentage of countries that were classified as following power law or exponential dynamics. We can see that for the countries that are just a few days behind Italy, 100% of them belong to the power law group. As the lag time increases, indicating an earlier stage of the epidemic, more and more countries exhibit exponential growth (*p* < 10^−4^).

**Figure 6:**
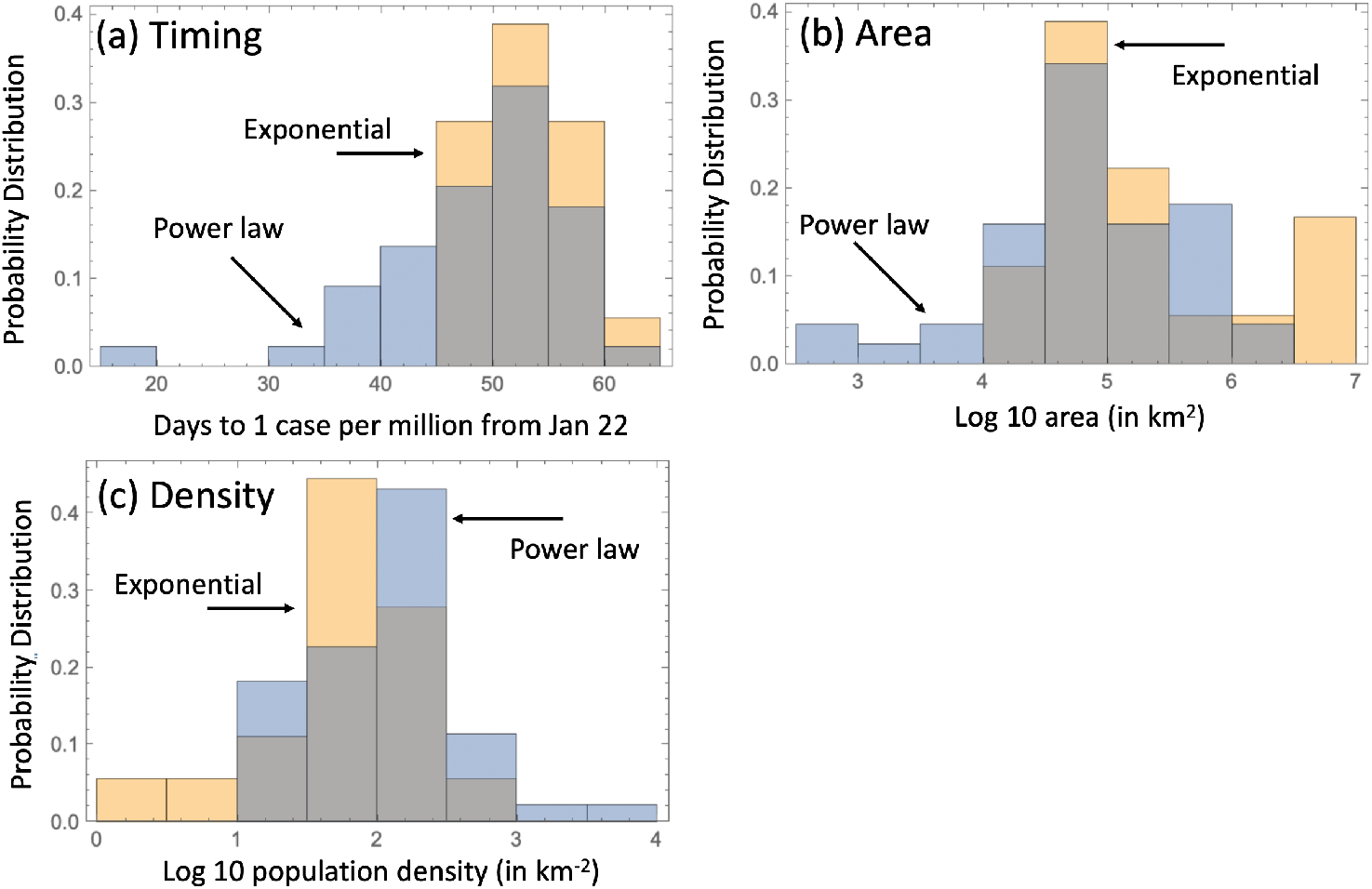
Comparison of the two classes of infection spread. (a) The timing of the infection: the distribution of time of reaching 1 case per million (counting from Jan 22), for the exponential and power law classes. The means are 48 days for the power law and 52 days for the exponential set (*p* = 0.035 by T Test). (b) Country size: the area of countries for the exponential and power law classes. The means are about 2.3 × 10^5^ km^2^ for the power law and 1.7×10^6^ km^2^ for the exponential, *p* = 0.018 by T test. (c) Country density: the means are about 374 people per km^2^ for the power law and 98 people per km^2^ for the exponential law, *p* = 0.035 by T test.

**Figure 7:**
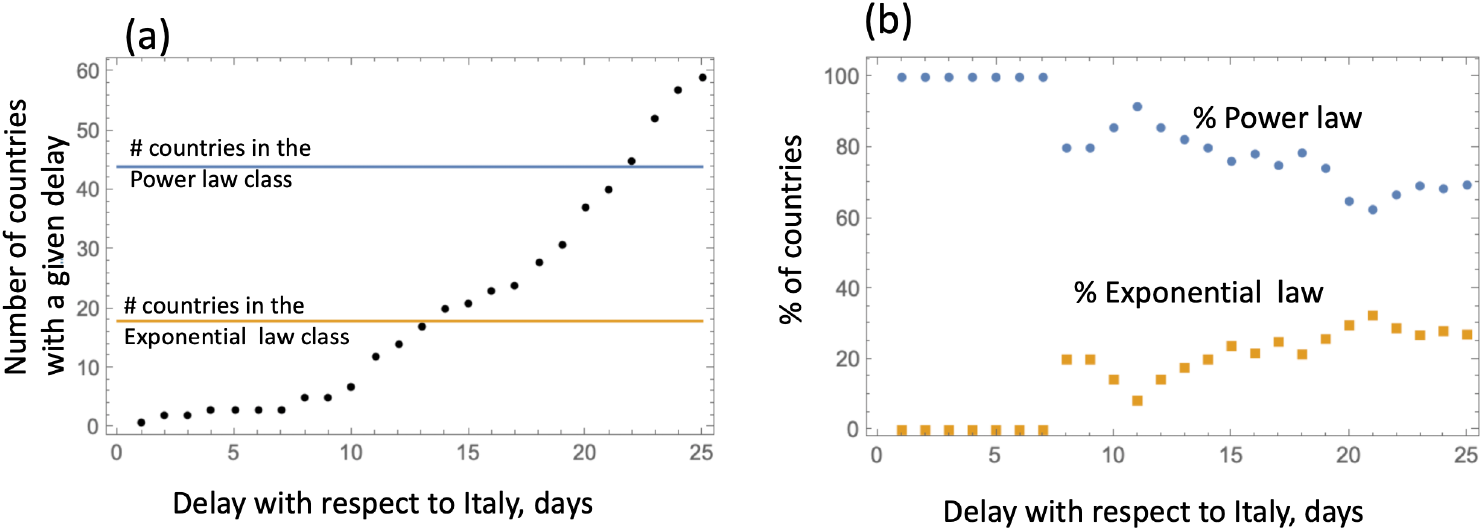
Temporal development of infection. The “age” of the epidemic is measured by the days of delay with respect to Italy. (a) The number of countries for each value of the time-lag with respect to Italy. The number of countries in the two growth law classes is shown for comparison. (b) The percentage of countries with a given delay that belong to the Power law group and to the Exponential law group. The trend that the percentage of exponential growth increases with the time lag (that is, decreases with the epidemic “age”) is significant (*p*< 10^−4^ by linear fitting).

### 3.4 Growth laws in relation to size and density of countries

Figure 6(b) shows the difference between countries with power law and exponential growth in terms of their area. We find that the exponentially growing infection class is associated with larger countries (mean area of about 1.7 × 10^6^ km^2^) compared to the power law class (mean area about 2.3 × 10^5^ km^2^, *p* = 0.018 by T test). Similarly, exponential epidemic spread tends to correlate with lower density countries (figure 6(c)). It is possible that it takes longer for a larger country of lower density to transition to a power law growth. Below, we provide a possible explanation of this correlation in the context of metapopulation modeling.

### 3.5 A metapopulation model can reproduce key trends in data

Our results can be interpreted in the context of a minimally parameterized metapopulation model, see figure 8. Assume that within a local deme (such as a local community), people interact with each other, resulting in mass action dynamics. For the infection to spread further, however, people have to enter other demes, and seed the infection there.

**Figure 8:**
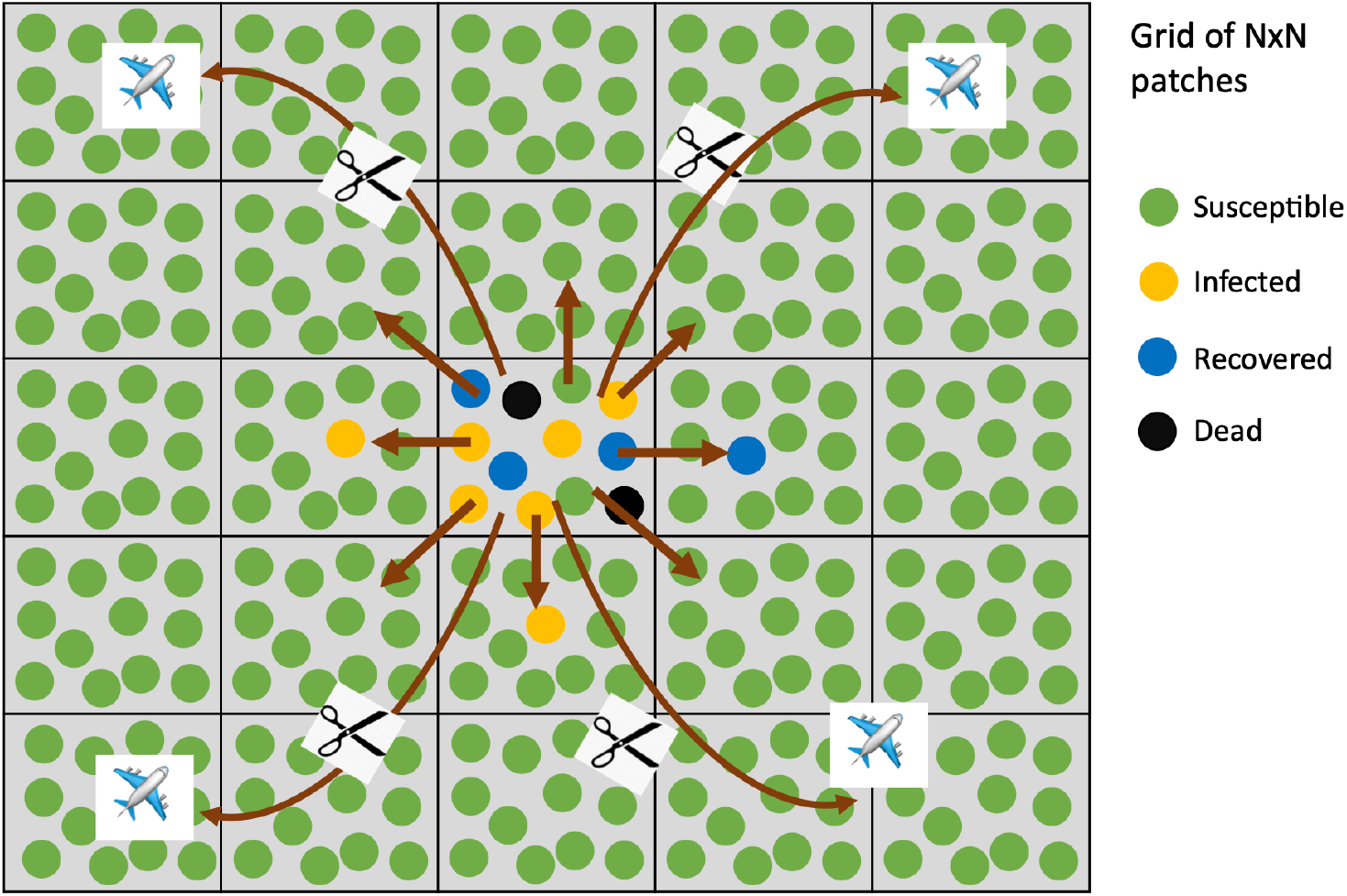
The concept of partial social distancing measures and the metapopulation model. There is a grid of *N* × *N* patches. Within each patch (which represents a local community), deterministic SIR dynamics are assumed (complete mixing). Infection can also spread by contact (mixing) with neighboring patches (demes). Global infection transfer is also possible, e.g. by air travel within the country and outside, but this is disrupted by partial social distancing measures. Equations (1-4) correspond to the situation where long-haul interactions are not prresent. This is what we implemented by simulations.

We have performed computer simulations of such a model to explore outcomes. The model is a two-dimensional metapopulation consisting of *N* × *N* patches. In each patch, *i*, the infection dynamics are given by a set of ordinary differential equations (ODEs) that take into account the population of susceptible (*S_i_*), infected (*I_i_*), recovered (*R_i_*),

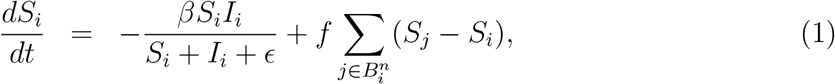

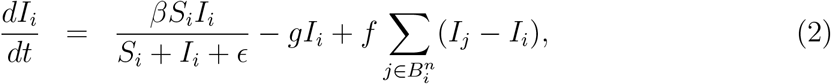

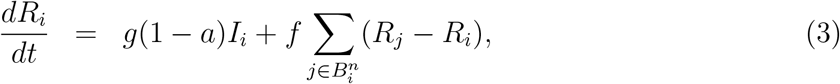

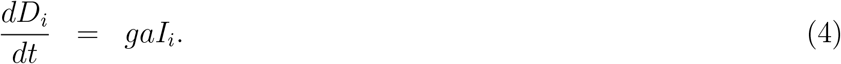

Here, infection is described by a frequency-dependent infection term [14], characterized but the rate constant *β* and a saturation constant ε. Infected individuals die with a rate *ga* and recover with a rate *g*(1 − *a*). The migration terms include the outward migration to n neighbors and an inward migration from all the n neighboring demes that belong to neighborhood 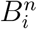 of deme *i*. The migration rate is denoted by f and we assume that each patch has eight direct neighbors, i.e. *n* = 8. The boundary demes are characterized by fewer inward/outward migrations (that is, they have smaller neighborhood sets).

Using this model, we track the predicted dynamics for *I* + *R* + *D* over time, which represents the cumulative infection case counts. In a first scenario, we start the computer simulations with a small amount of infected individuals in a single patch, located in the center of the grid. All other patches contain only susceptible individuals. The resulting dynamics are shown in figure 9. We observe an initial exponential phase of infection spread, followed by a transition to a power-law spread. The spread is initially exponential, because within a single patch (the first patch), the dynamics are governed by well mixed populations. As the infection spreads to other patches by migration, the overall infection spread starts to be governed by spatial dynamics, which explains the transition to the power law behavior (see [15, 16] for the mathematical treatment of epidemic spread in 2D). The key is the difference between the time scale of local spread and the time-scale of global mixing.

**Figure 9:**
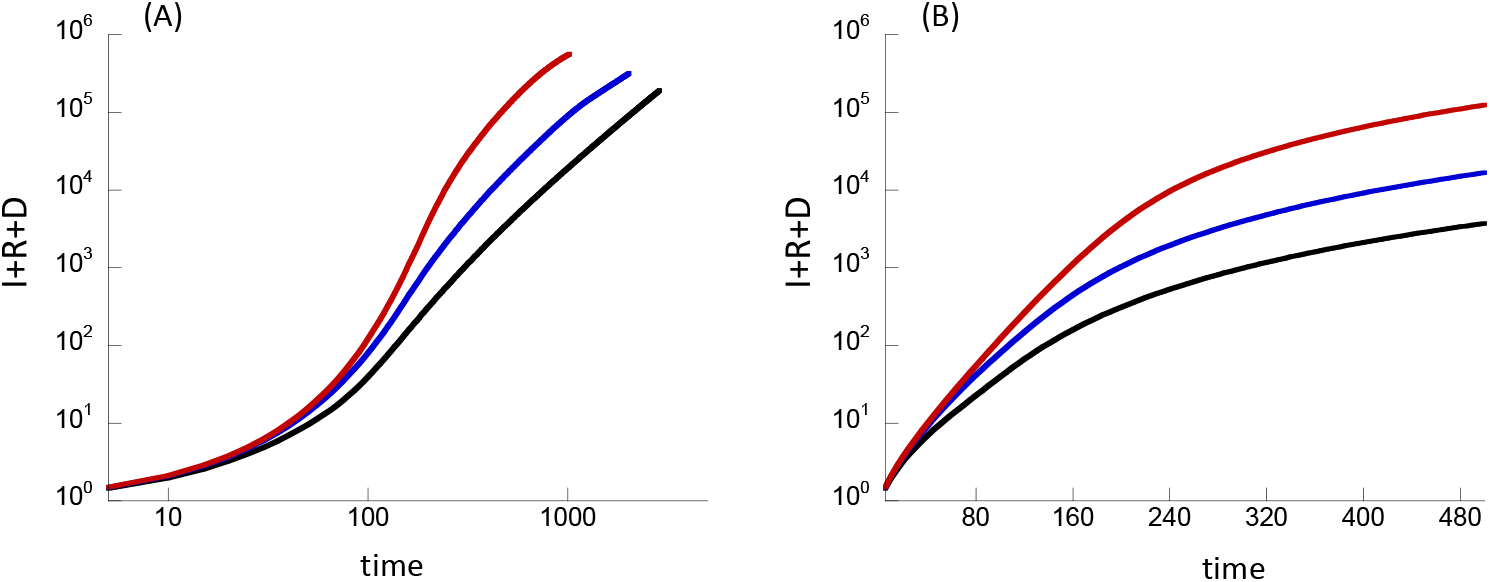
Results from implementing the metapopulation model, equations (1-4). The total number of cases (given by *I*+*R*+*D*) is plotted as a function of time, on a log log scale (a) and a log scale (b). The black line shows the dynamics where the simulation starts with 1/10 of individuals infected in a single patch in the middle. The blue line corresponds to the initial condition where 1/50 of the individuals are infected in 5 randomly chosen patches. The red line shows the consequence of 1/500 of the individuals infected in 50 randomly chosen patches. The rest of the parameters are *S* = 10,*R* = 0, *D* = 0 initially in all patches, *β* = 0.1, *g* = 0.05, *f* = 0.001, *a* = 0.01,∊ = 1, *N* = 300.

Next we assumed that instead of starting with infecteds only being present in a single patch, a small amount of infected individuals are initially present in more than one patch around the same time. This could correspond to larger countries, in which the infection is simultaneously seeded in multiple areas (e.g. due to travel from other places). Now, we observe overall growth dynamics that are are more exponential-like. The length of the predicted exponential phase becomes longer the more patches are initially seeded. The reason is that with more initial seeding events, the importance of spatial spread is de-emphasized.

In summary, the metapopulation model can predict an array of growth patterns where an exponential phase of varying length is followed by a transition to power law, depending on the initial conditions of the simulation.

## 4 Discussion and Conclusions

In this paper, we analyzed data that document the cumulative COVID19 case counts over time in a large number of countries around the world, and examined the laws according to which the infection spreads. This suggests that although the initial phase of the spread may be exponential, the longer term dynamics (extending to March 28, 2020) tend to be governed by a power law. The analysis indicates that the countries that displayed clear evidence for exponential growth were in a relatively early phase of the epidemic, and that countries that were further along in the epidemic converged to a power law behavior.

These observations were interpreted by computer simulations of a metapopulation model that takes into account both local spread and spread across geographical space. This model predicts an initial exponential phase (due to local transmission events driving the dynamics), followed by a transition to a power law (once spatial dynamics significantly drive spread). The duration of the exponential phase is determined by the number of patches that are initially seeded with the infection. If the infection originates in a single location (patch), the exponential phase is likely not very pronounced, and most of the growth curve is predicted to follow a power law. If the infection is seeded simultaneously or nearly simultaneously in multiple locations, the duration of the exponential phase becomes longer. This might explain why countries with larger areas show more pronounced exponential growth, as this makes multiple seeding events due to travel more likely. Interestingly, it has been shown that in China outside the Hubei province, COVID19 cases initially grew relatively fast, and this was shown to correlate with human movement out of Wuhan into the affected areas [17], indicating multiple seeding events. These model predictions could also imply a more extensive exponential phase of infection spread during second waves, after non-pharmaceutical interventions have been relaxed. The reason is that once the infection has already propagated through the community before the interventions, it is already extensively seeded across most areas, resulting in more pronounced exponential growth and less pronounced growth that is governed by spatial spread.

A better understanding of the laws according to which COVID19 spreads through populations is also of practical importance. (i) Projections and forecasts made under the assumption of exponential growth lead to significantly faster virus spread compared to those assuming power-law growth, as shown in the Supplemental Section E. (ii) As COVD19 outbreaks in a given region or country unfold, an eventual deviation from exponential growth and an estimated longer doubling time does not indicate that the infection is being controlled, for example by non-pharmaceutical interventions. Power law growth is characterized by a naturally decreasing “doubling time” (even in the absence of interventions), and the repeated estimation of doubling times are not meaningful in this case. To establish that interventions flatten the curve, it has to be demonstrated that growth either transitions to a lower power, or that it deviates entirely from a power law, which has occurred in many countries after the time period under consideration here. (iii) Similarly, the power growth laws obtained in our analysis have implications for the estimation of the effective reproduction number, see e.g. [18]. As the infection continues to spread according to power laws, the effective reproduction number becomes lower over time. Again, however, this is not necessarily the result of interventions, but a natural consequence of the power law. This is important to keep in mind when estimating the reproductive potential of SARS-CoV-2.

Alternative explanations can be invoked to account for sub-exponential growth patterns. Data indicate that in China, the implementation of non-pharmaceutical interventions can drive sub-exponential growth by depleting the pool of susceptible individuals over time through social distancing [9], and this can lead to a temporary phase of power law growth. An initial fast virus spread phase, followed by a slow-down of spread, can also come about if the initial phase of spread is driven mostly by immigration of infected individuals from other geographical areas, while subsequent community spread continues with a slower rate [17]. While these mechanisms are certainly plausible, it would be expected that growth patterns are different depending on the timing at which interventions are implemented, the strength at which they are implemented, or depending on the magnitude of infection seeding by immigration. Yet, we observe remarkably consistent growth laws in a number of different countries, in which policies in response to the outbreak varied, and in which the magnitude of initial seeding events likely differed. This points towards the possibility that the power law growth dynamics are an intrinsic feature of COVID19 spread through human populations, and that they are not externally imposed.

As with any data and modeling studies, it is important to note that results can depend on assumptions and methodologies. These are clearly spelt our here. One of the larger challenges we faced in the data analysis is the lack of knowledge at what time the infection was initiated in the individual countries. This information is not available. The time frame in turn influences the fit of the power law to the data, which we have addressed with our time shifting methodology. If further information becomes available about the time when infections are estimated to have originated in the individual countries, the methodology can be updated. Genetic studies could provide valuable data in this respect.

Another limitation of the data interpretation is the degree to which different countries test for SARS-CoV-2. If some countries test less than others, they will appear to be at an earlier stage of the outbreak than is true. This type of uncertainty however does not change the central finding that the long term dynamics of COVID19 cases in different countries follow a power law, after an initial stage of exponential growth. Another testing-related problem could be if the number of tests in a given country changes over time. Typically, the level of testing has increased over time (Supplemental Section C), but we argue that it is unlikely that an increasing number of tests over time would invalidate our finding that instead of growing exponentially, the number of cases grow according to a power law. Increased testing over time would accelerate the growth rate and could potentially make the growth curve look more exponential, meaning that the power laws we found are not likely to be an artifact of varying testing levels. In fact, a fast growth in the number of tests could shift some countries where the true number of cases grew as a power law to an “exponential” category, because of the accelerating effect of the testing. We also considered COVID19-related deaths as a measure of disease spread (Supplemental Section D), and found that our conclusions remained robust. Deaths are less likely to depend on testing numbers, but are connected with their own set of challenges. Different countries use different case definitions, testing criteria often change even in a given country as the local epidemic progresses, and different definitions of what constitutes a COVID19 death are used in different jurisdictions, and even at different stages of any given outbreak. While each individual measure of COVID19 spread is connected to problems that make it difficult to interpret the data, the consistent patterns that we found across those different measures strengthen our conclusions.

## Data Availability

All data that are analyzed in this paper have been obtained from an online repository. The data are freely available for download at this site, and the web location is specified in the paper.

https://datahub.io/core/covid-19#data-cli

## Acknowledgements

Support of grant NSF DMS 1662146/1662096 is gratefully acknowledged.

## References

[1] Velavan TP, Meyer CG. The COVID-19 epidemic. Trop Med Int Health. 2020;25(3):278–280.

[2] Rothan HA, Byrareddy SN. The epidemiology and pathogenesis of coronavirus disease (COVID-19) outbreak. Journal of Autoimmunity. 2020;p. 102433.

[3] Zhou F, Yu T, Du R, Fan G, Liu Y, Liu Z, et al. Clinical course and risk factors for mortality of adult inpatients with COVID-19 in Wuhan, China: a retrospective cohort study. The Lancet. 2020;.

[4] Emanuel EJ, Persad G, Upshur R, Thome B, Parker M, Glickman A, et al. Fair allocation of scarce medical resources in the time of Covid-19. Mass Medical Soc; 2020.

[5] Ferguson NM, Laydon D, Nedjati-Gilani G, Imai N, Ainslie K, Baguelin M, et al. Impact of non-pharmaceutical interventions (NPIs) to reduce COVID-19 mortality and healthcare demand. Imperial College, London DOI: https://doiorg/1025561/77482. 2020;.

[6] Remuzzi A, Remuzzi G. COVID-19 and Italy: what next? The Lancet. 2020;.

[7] Li Y, Liang M, Yin X, Liu X, Hao M, Hu Z, et al. COVID-19 Epidemic Outside China: 34 Founders and Exponential Growth. medRxiv. 2020;.

[8] Anderson RM, May RM. Infectious diseases of humans: dynamics and control. Oxford university press; 1992.

[9] Maier BF, Brockmann D. Effective containment explains subexponential growth in recent confirmed COVID-19 cases in China. Science. 2020;368(6492):742–746.

[10] Novel Coronavirus 2019, by Johns Hopkins University Center for Systems Science and Engineering; 2020. https://datahub.io/core/covid-19#data-cli.

[11] Our World in Data; 2020. https://ourworldindata.org/grapher/full-list-total-tests-for-covid-19.

[12] Dong E, Du H, Gardner L. An interactive web-based dashboard to track COVID-19 in real time. The Lancet infectious diseases. 2020;20(5):533–534.

[13] Li M, Chen J, Deng Y. Scaling features in the spreading of COVID-19. arXiv preprint arXiv:200209199. 2020;.

[14] McCallum H, Barlow N, Hone J. How should pathogen transmission be modelled? Trends in ecology & evolution. 2001;16(6):295–300.

[15] Cox J, Durrett R. Limit theorems for the spread of epidemics and forest fires. Stochastic processes and their applications. 1988;30(2):171–191.

[16] Durrett R. Spatial epidemic models. In: Mollison D, editor. Epidemic models: their structure and relation to data. Cambridge: Cambridge Univ. Press; 1995. p. 187–201.

[17] Kraemer MU, Yang CH, Gutierrez B, Wu CH, Klein B, Pigott DM, et al. The effect of human mobility and control measures on the COVID-19 epidemic in China. Science. 2020;368(6490):493–497.

[18] Wu JT, Leung K, Leung GM. Nowcasting and forecasting the potential domestic and international spread of the 2019-nCoV outbreak originating in Wuhan, China: a modelling study. The Lancet. 2020;395(10225):689–697.

[19] Novel Coronavirus 2019; 2020. https://datahub.io/core/covid-19#data-cli.

[20] Dipartimento della Protezione Civile COVID-19 Italia-Monitoraggio della situazione; 2020. http://opendatadpc.maps.arcgis.com/apps/opsdashboard/index.html#/b0c68bce2cce478eaac82fe38d4138b1.

